# Comparison of epicardial adipose tissue volume quantification between cardiac and chest computed tomography scans

**DOI:** 10.1101/2020.11.26.20239053

**Authors:** Lingyu Xu, Yuancheng Xu, Stanislau Hrybouski, D Ian Paterson, Richard B. Thompson, Zhiyang Li, Yulong Lan, Craig Butler

**Affiliations:** Department of Cardiology, Mackenzie Health Science Centre, Faculty of Medicine & Dentistry, University of Alberta, Edmonton, Alberta, Canada; Department of Biomedical Engineering, University of Alberta, Edmonton, Canada; Department of Urology, The University of Hong Kong-Shenzhen Hospital, Shenzhen, Guangdong, China; Neuroscience and Mental Health Institute, University of Alberta, Edmonton, Alberta, Canada; Department of General Surgery, the Second Affiliated Hospital of Shantou University Medical College, Shantou, Guangdong, China; Department of Cardiology, the Second Affiliated Hospital of Shantou University Medical College, Shantou, Guangdong, China

**Keywords:** Epicardial adipose tissue, Chest Computed tomography, Tube energy, Contrast enhancement, Slice thickness

## Abstract

**Background:** This study investigated accuracy and consistency of epicardial adipose tissue (EAT) quantification in chest computed tomography (CT) scans.

**Methods and results:** EAT volume was quantified semi-automatically using a standard Hounsfield unit threshold (-190U, -30) in three independent cohorts: (1) Cohort 1 (*N* = 30) consisted of paired 120 KV cardiac non-contrast CT (NCCT) and 120 KV chest NCCT; (2) Cohort 2 (*N* = 20) consisted of paired 120 KV cardiac NCCT and 100 KV chest NCCT; (3) Cohort 3 (*N* = 20) consisted of paired chest NCCT and chest contrast-enhanced CT (CECT) datasets. Images were reconstructed with the slice thicknesses of 1.25 mm and 5 mm in the chest CT datasets, and 3 mm in the cardiac NCCT datasets. In Cohort 1, the chest NCCT-1.25 mm EAT volume was similar to the cardiac NCCT EAT volume, whilst chest NCCT-5 mm underestimated the EAT volume by 7.0%. In Cohort 2, 100 KV chest NCCT-1.25mm and -5 mm EAT volumes were 9.7% and 6.4% larger than corresponding 120 KV cardiac NCCT EAT volumes. In Cohort 3, the chest CECT dataset underestimated EAT volumes by ∼25%, relative to chest NCCT datasets. All chest CT-derived EAT volumes were strongly correlated with their cardiac CT counterparts.

**Conclusions:** The chest NCCT-1.25 mm EAT volume with the 120 KV tube energy produced EAT volumes that are comparable to cardiac NCCT. All chest CT EAT volumes were strongly correlated with EAT volumes obtained from cardiac CT, if imaging protocol is consistently applied to all participants.

## 1. Introduction

The epicardial adipose tissue (EAT) – located between the outer wall of the myocardium and the visceral pericardium – has proatherogenic effect on the coronary arteries via secretion of pro-inflammatory cytokines.^1, 2^ Traditionally, EAT volumes have been measured using cardiovascular non-contrast computed tomography (cardiac NCCT) images.^1, 3, 4^ More recently, EAT volumes have been reported from chest CT images^5-8^. Chest CT scans provide an appealing alternative to cardiac NCCT because of their widespread use in clinical practice. ^9^ Examining EAT volumes using chest CT may enable early stratification of cardiovascular risk for patients undergoing clinical chest CT scans.^8, 10^ Currently, cardiac NCCT acquisitions with electrocardiogram (ECG) gating, tube energy of 120 KV, and a slice thickness of 3 mm are the standard for quantifying EAT volume.^3, 11, 12^ The effects of chest CT acquisition and reconstruction parameters – i.e., presence vs. absence of ECG gating or contrast agents, differences in tube energy and slice thickness, relative to cardiac CT scans – on calculated EAT volumes are still poorly understood.

Chest CT does not typically use ECG gating, making the heart and EAT pool susceptible to artifacts from cardiac motion. Furthermore, without ECG gating, chest CT, unlike cardiac CT, is unable to acquire images at fixed points of the cardiac cycle.^13^ Tube energy substantially affects tissue radiodensity in the CT images.^14^ Given that EAT volume quantification is based on a radiodensity range(e.g., -190 to -30 HU), ^3, 4, 7, 15-17^ understanding impacts of tube energy on EAT volumes derived from chest CT scans is vital for measurement consistency with cardiac CT scans. The reconstructed slice thickness is commonly 3 mm in the cardiac NCCT image, but this is not a routine in chest CT image, and the slice thicknesses in clinical chest CT scans can vary substantially, with 1 mm, 1.25 mm, 2.5 mm and 5 mm as the most routinely used.^5, 8, 18, 19^ Thus, the effects of chest CT slice thickness on EAT volumes need to be investigated further. Finally, contrast enhancement has been shown to underestimate EAT volume derived from cardiac contrast-enhanced CT images (cardiac CECT) compared to those derived from cardiac NCCT images using the same radiodensity thresholds.^11, 17^ To date, no study has investigated the effect of contrast agents on chest CT-derived EAT.

This study aims to investigate the feasibility of studying EAT volumes using a number of commonly acquired chest CT datasets. Chest CT EAT volumes were then compared to those that were obtained using the gold standard setup for EAT quantification – 120 KV cardiac NCCT. Additionally, we evaluated the effect of contrast enhancement on EAT volume measurement in chest CT.

## 2. Methods

### 2.1 Patient recruitment

We retrospectively identified patients who underwent paired chest NCCT and cardiac NCCT scans between 2016 and 2019 from the University of Hong Kong-Shenzhen Hospital clinical database. To minimize the effect of time on EAT volume, time interval between paired CT scans was restricted such that the two scans were acquired no more than two weeks apart. For chest NCCT vs. chest CECT comparisons, both datasets were acquired in the same session in January 2019. This study was approved by the local human research ethics committee.

The imaging protocol is summarized in Table 1. Two slice thicknesses of 1.25 mm and 5.0 mm were used separately for chest CT image reconstruction in all patients and used for comparisons in all cohorts. Cohort 1 consisted of 30 patients who underwent paired cardiac and chest NCCT scans (both with tube energy 120KV). Cohort 2 consisted of 20 patients who underwent paired cardiac NCCT scans (120 KV) and chest NCCT (100 KV). Lastly, the effects of contrast enhancement on EAT volumes were quantified using paired chest CT datasets (i.e., chest NCCT vs chest CECT; Cohort 3, N = 20).

**Table 1.** CT acquisition and reconstruction parameters in three cohorts

|  | Cohort 1 (N = 30) |  | Cohort 2 (N = 20) |  | Cohort 3 (N = 20) |  |
| --- | --- | --- | --- | --- | --- | --- |
| Paired CT dataset | Cardiac NCCT | Chest NCCT | Cardiac NCCT | Chest NCCT | Chest NCCT | Chest CECT |
| ECG gate | Yes | No | Yes | No | No | No |
| Tube energy (KV) | 120 | 120 | 120 | 100 | 120 | 120 |
| Contrast given | No | No | No | No | No | Yes |
| Collimation (mm) | 128×0.6 | 128×0.625 | 128 × 0.6 | 128×0.625 | 128×0.625 | 128×0.625 |
| Gantry rotation time (ms) | 330 | 350 | 330 | 350 | 350 | 350 |
| Pitch | 0.24 | 1.2 | 0.24 | 1.2 | 1.2 | 1.2 |
| Reconstructed Slice thickness (mm) | 3 | 1.25 & 5 | 3 | 1.25 & 5 | 1.25 & 5 | 1.25 & 5 |

Abbreviation: ECG= electrocardiogram, NCCT = non-contrast computed tomography, CECT = contrast-enhanced computed tomography.

### 2.2 Cardiovascular CT protocol

Our cardiac NCCT images were acquired using a 64 detector-row CT scanner (Somatom Definition AS, Siemens, Germany) with the following parameters: retrospective ECG gating, tube energy = 120 KV, collimation = 128 × 0.6 mm, gantry rotation time = 330 ms, pitch = 0.24. Z-axis coverage extended from the pulmonary artery bifurcation to the ventricular apex. Reconstruction was performed at 70% of the RR wave, and images were reconstructed with 3-mm slice thickness and 1.5 mm interslice gap using soft-tissue convolution kernel (B35f). To maintain heart rate at or below 70 bpm, patients were pre-treated with oral metoprolol as necessary (see Table 1).

### 2.3 Chest CT protocol

Chest CT images were acquired using a 64 detector-row CT scanner (LightSpeed VCT, GE, USA) with following parameters: collimation = 128 × 0.625 mm, gantry rotation time = 350 ms, pitch = 1.2. The tube energy was 120 KV in Cohorts 1 and 3, and 100 KV in Cohort 2. Images were reconstructed as 1.25-mm thick slices (+1-mm interslice gap) and 5-mm thick slices (+5-mm interslice gap), respectively. A soft-tissue convolution kernel of B31f was used. Participants in Cohort 3 first underwent chest NCCT image acquisition and then, to acquire the chest CECT image, they were intravenously administered a bolus of contrast medium (Iopromide 370, Bayer Schering Pharma AG; dosage = 1 ml/kg body weight, rate = 3 ml/s) followed by a flush of normal saline (30 ml). Coverage in all chest CT datasets extended from the thoracic inlet to the upper abdomen, at a minimum.

### 2.4 Epicardial adipose tissue volume quantification

The EAT volume was quantified using freely-available a dedicated image analysis software (ITK-SNAP version 3.6.0).^20^ A widely used radiodensity threshold (−190, -30) HU^3, 4, 7, 15-17^ map was created in-house using in-house MATLAB code and was applied to all cardiac and chest CT images. Longitudinally, EAT quantification began from the bifurcation of the pulmonary artery and ended at the level of the left ventricular apex. The contour of the pericardium was manually traced on all slices. Finally, the EAT volume and radiodensity were automatically computed (Figure 1).

**Figure 1.**
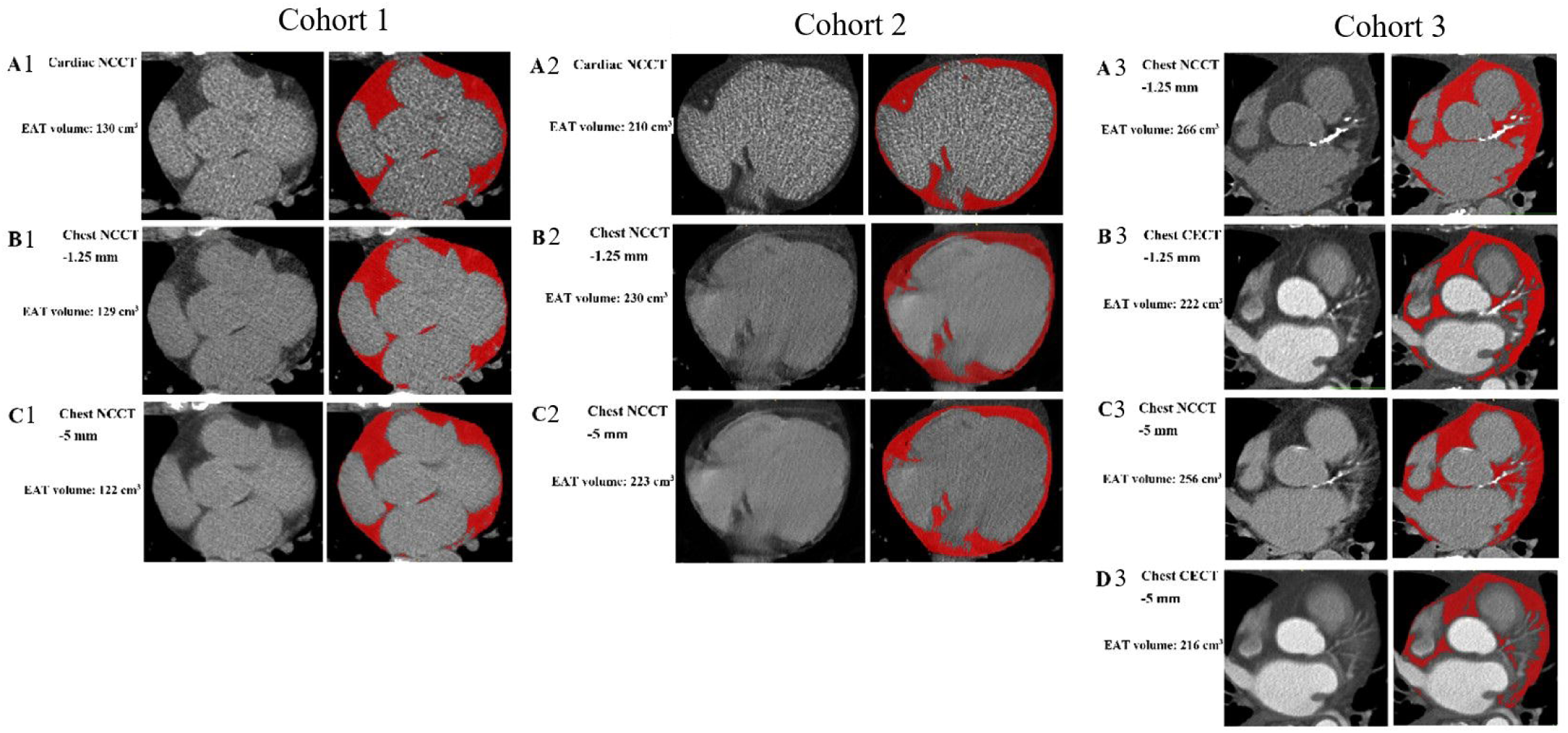
Examples of measuring and comparing EAT volumes at the same slice in paired CT datasets of three cohorts. Each cohort includes CT images in left panel and EAT highlighted in red in the right panel which was semi-automatically quantified using radiodensity threshold of - 190 HU, -30 HU.1A): Cohort 1, cardiac NCCT (120 KV) *vs*. chest NCCT-1.25 mm (120 KV) *vs*. chest NCCT-5mm (120 KV); 1B): Cohort 2, cardiac NCCT (120 KV) *vs*. chest NCCT-1.25 mm (100 KV) *vs*. chest NCCT-5mm (100 KV); 1C): chest NCCT-1.25 mm (120 KV) *vs*. chest CECT-1.25 mm (120 KV), and chest NCCT-5mm (120 KV) *vs*. chest CECT-5mm (120 KV). Abbreviation: Abbreviation: EAT = epicardial adipose tissue, CT= computed tomography, NCCT = non-contrast computed tomography, CECT = contrast-enhanced computed tomography, ECG= electrocardiogram, vs. = versus, HU = Hounsfield unit.

### 2.5 Statistics

Statistical analyses were performed in STATA (Version 16.0; StataCorp LP, College Station, Texas, USA). Continuous variables were presented as mean ± standard deviation unless otherwise indicated. Categorical variables were expressed as frequencies and percentages. Paired t-tests were used to evaluate the statistical significance of EAT volumes. One-way analysis of variance and Chi-squared tests were used to compare sample characteristics among cohorts. Shapiro-Wilk tests were used to test for normality violations. Bland-Altman analyses were used for assessing consistency of EAT measurements from different CT scans. *P-*values less than 0.05 were used as a measure of significance in all tests.

## 3. Results

The demographic data and disease history of three cohorts are summarized in Table 2. Mean age for the entire sample was 63 years with 64% males. Three cohorts had similar body mass index and disease history. The time interval between chest CT and cardiac CT scans (in Cohorts 1 and 2) was 6 ± 4 days.

**Table 2.** Basic characteristics in three cohorts

|  | Overall<br>(N = 70) | Cohort 1<br>(N = 30) | Cohort 2<br>(N = 20) | Cohort 3<br>(N = 20) | P-value |
| --- | --- | --- | --- | --- | --- |
| Age (years) | 63±11 | 64±12 | 58±9 | 68±9 | 0.02 |
| Male (%) | 45(64%) | 19(63%) | 13(65%) | 14(70%) | 0.80 |
| Body mass index (kg/m<sup>2</sup>) | 24.8±3.7 | 25.0±3.4 | 24.7±4.0 | 24.6±3.9 | 0.94 |
| Coronary artery disease | 11(16%) | 3(10%) | 6(30%) | 2(10%) | 0.15 |
| Hypertension | 24(34%) | 10(33%) | 6(30%) | 8(40%) | 0.79 |
| Diabetes mellitus | 8(11%) | 3(10%) | 5(25%) | 0(0%) | 0.03 |
| Smoking | 12(17%) | 4(13.3%) | 2(10%) | 6(30%) | 0.29 |
| Hyperlipidemia | 11(16%) | 3(10%) | 6(30%) | 2(10%) | 0.15 |
| Chronic obstructive<br>pulmonary disease | 14(20%) | 5(17%) | 2(10%) | 7(35%) | 0.15 |

### 3.1 EAT volume in Cohort 1

The comparison of EAT volume quantification between the gold standard (cardiac NCCT image, 120KV, 3-mm slice thickness) and 120 KV chest NCCT image (1.25-mm & 5-mm slice thickness) is summarized in Table 3 & Figure 2. Compared to standard cardiac NCCT EAT volumes, the chest NCCT-1.25 mm EAT volumes were similar (133.7±44.7 *vs*.134.2±43.3 cm^3^, Δ%=-0.8±3.8%, *p* = 0.46), and the Bland-Altman analysis also demonstrated an insignificant difference of 0.6 (−1.0, 2.2) cm^3^. However, 5-mm chest NCCT produced that were consistently lower than those based on the cardiac NCCT scans (125.0±40.6 *vs*.134.2±43.3 cm^3^, Δ%=-7.0±2.5%, *p* <0.001). The EAT radiodensity for the chest NCCT-1.25 mm scans (−78.8±8.8 HU) was similar to that of the cardiac NCCT EAT scans (−78.2±5.8 HU), while the EAT radiodensity of the chest NCCT-5 mm scans (−74.2±7.3 HU) was significantly higher (i.e., less negative) than that of the cardiac NCCT EAT scans. Both chest NCCT EAT volumes were highly correlated with cardiac NCCT EAT volumes (See Figure 2). The chest NCCT-1.25 mm EAT volumes were higher than the chest CT-5 mm EAT volumes (133.7±44.7 cm^3^ *vs*. 125.0±40.6 cm^3^, Δ%=-6.1±3.3%, *p* < 0.001, see Table 3 & Figure 3A).

**Table 3.** Comparison of EAT volume between cardiac NCCT and chest NCCT image at the same tube energy (120 KV) in Cohort 1 (N=30)

|  | Cardiac NCCT | Chest NCCT-1.25mm | Chest NCCT-5mm |
| --- | --- | --- | --- |
| EAT volume (cm<sup>3</sup>) | 134.2±43.3 | 133.7±44.7 | 125.0±40.6 |
| ΔVolume, cm<sup>3</sup> (%)<br>(vs. Cardiac NCCT) | 0 | -0.6±4.4<br>(-0.8±3.8%) | -9.2±4.5<br>(-7.0±2.5%) |
| P-value 1 (vs. Cardiac<br>NCCT EAT volume) | 1.0 | 0.46 | <0.001 |
| EAT radiodensity (HU) | -78.2±5.8 | -78.9±8.8 | -74.2±7.3 |
| P-value 2 (vs. Cardiac<br>NCCT EAT radiodensity) |  | 0.21 | 0.002 |

Abbreviation: EAT = epicardial adipose tissue, ECG= electrocardiogram, NCCT = non-contrast computed tomography, vs. = versus, HU = Hounsfield unit.

**Figure 2.**
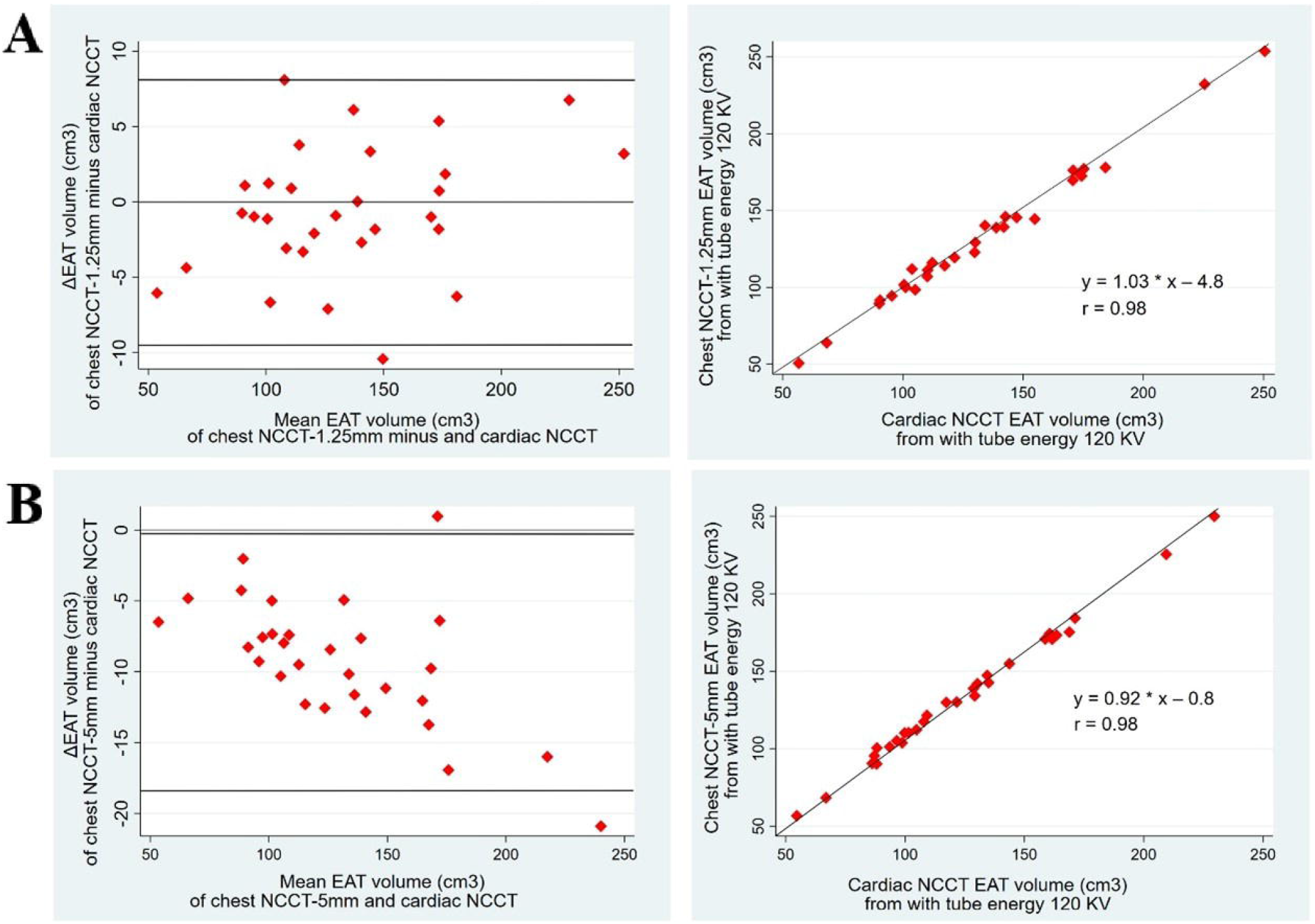
The relationship between cardiac NCCT EAT volumes and chest NCCT EAT volumes at the same tube energy (120 KV) in Cohort 1. EAT volume was semi-automatically quantified using radiodensity threshold of -190 HU, -30 HU. Bland-Altman plots (Left panel) and scatter plots (Right panel) of EAT volume between cardiac NCCT images (referent) and chest NCCT-1.25 mm images, and between cardiac NCCT images (referent) and chest NCCT-5 mm image, respectively. Abbreviation: see Figure 1.

**Figure 3.**
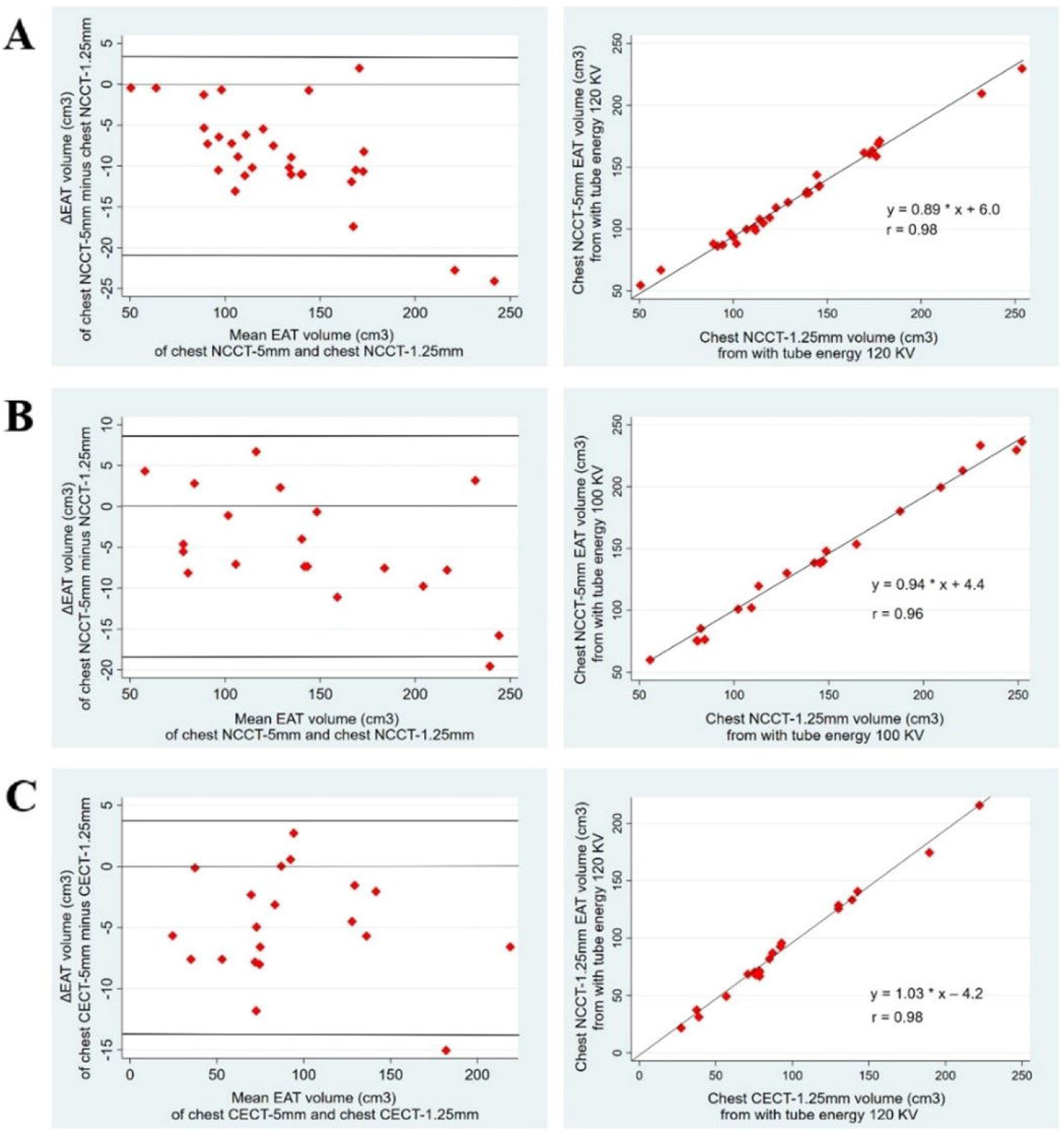
Effects of different slices thickness on EAT volume quantification in paired chest CT-1.25 mm and chest CT-5 mm images when other acquisition parameters keep fixed. Left panel: Bland-Altman plots (Left panel) and scatter plot (Right panel) of EAT volume between chest CT-1.25 mm images (referent) and chest CT-5 mm images both acquired without contrast enhancement at 120 KV in the upper panel, both acquired without contrast enhancement at 100 KV in the middle panel, and both acquired with contrast enhancement at 120 KV in the lower panel, respectively. Abbreviation: see Figure 1.

### 3.2 EAT volume in Cohort 2

The comparison of EAT volume quantification between the gold standard (cardiac NCCT image, 120KV, 3-mm slice thickness) and 100 KV chest NCCT image (1.25-mm & 5-mm slice thickness) is summarized in Table 4 & Figure 4. Using the tube energy of 100 KPV, chest NCCT-1.25 mm EAT volume was overestimated compared to the standard cardiac NCCT EAT volume using the tube energy of 120 KPV (146.6±60.6 *vs*. 133.0±52.4 cm^3^, Δ%=9.7±5.1%, *p* <0.001), and the Bland-Altman analysis demonstrated a systemic overestimation of 13.6 (8.7, 18.5) cm^3^. For the chest NCCT-5mm, our results showed that EAT volume (141.7±57.0 *vs*. 133.0±52.4 cm^3^, Δ%=6.4±3.3%, *p* <0.001) was overestimated by 8.7 (5.7, 11.7) cm^3^, on average. Both chest NCCT EAT volumes had strong correlations with cardiac NCCT EAT volumes (Figure 4). Furthermore, chest NCCT-1.25 mm EAT radiodensity (−84.2±6.6 HU) and chest NCCT-5 mm EAT radiodensity (−80.5±6.2 HU) were significantly lower (more negative) than the cardiac NCCT EAT radiodensity (−78.4±4.8 HU), both *p*s<0.05. The chest CT-1.25 mm EAT volumes were higher than the chest CT-5 mm EAT volumes (146.6±60.6 cm^3^ *vs*. 141.7±57.0 cm^3^, Δ%=-2.7±4.7%, *p* = 0.016, see Table 4 & Figure 3B)

**Table 4.**
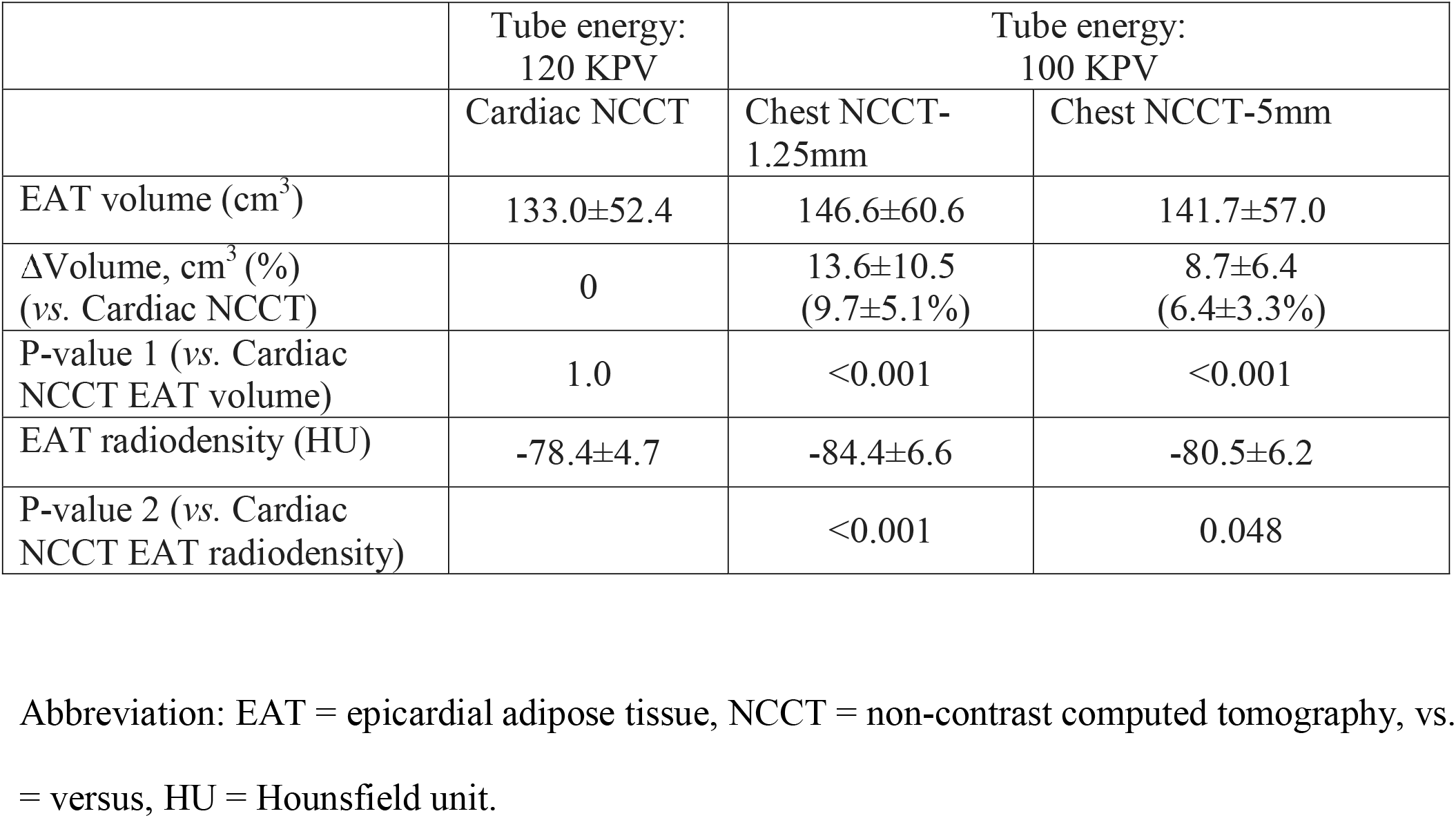
Comparison of EAT volume between standard cardiac NCCT and chest NCCT image with a lower tube energy (100KV) in Cohort 2 (N=20)

|  | Tube energy:<br>120 KPV | Tube energy:<br>100 KPV |  |
| --- | --- | --- | --- |
|  | Cardiac NCCT | Chest NCCT-1.25mm | Chest NCCT-5mm |
| EAT volume (cm<sup>3</sup>) | 133.0±52.4 | 146.6±60.6 | 141.7±57.0 |
| ΔVolume, cm<sup>3</sup> (%)<br>(vs. Cardiac NCCT) | 0 | 13.6±10.5<br>(9.7±5.1%) | 8.7±6.4<br>(6.4±3.3%) |
| P-value 1 (vs. Cardiac<br>NCCT EAT volume) | 1.0 | <0.001 | <0.001 |
| EAT radiodensity (HU) | -78.4±4.7 | -84.4±6.6 | -80.5±6.2 |
| P-value 2 (vs. Cardiac<br>NCCT EAT radiodensity) |  | <0.001 | 0.048 |

Abbreviation: EAT = epicardial adipose tissue, NCCT = non-contrast computed tomography, vs. = versus, HU = Hounsfield unit.

**Figure 4.**
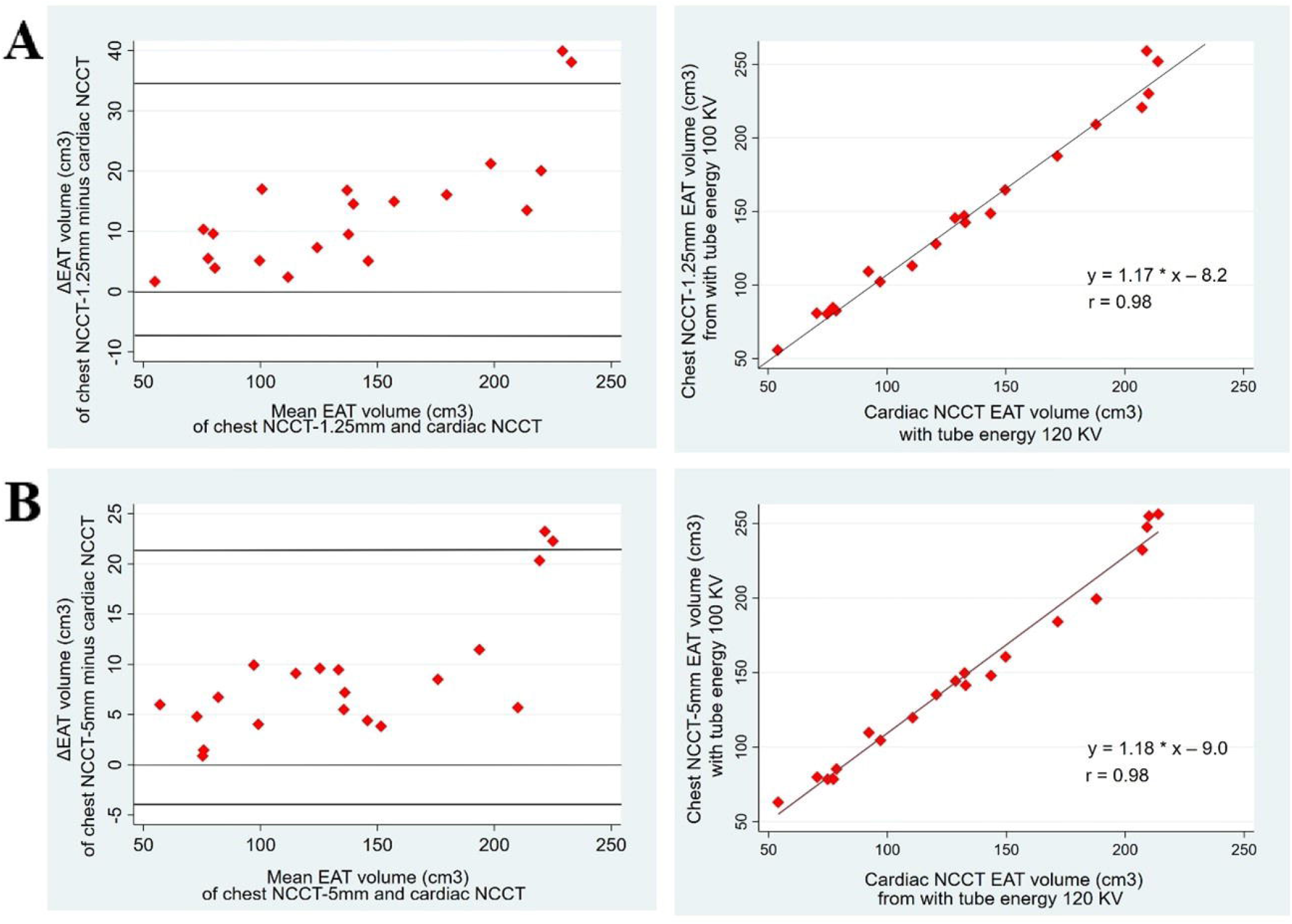
The relationship between cardiac NCCT EAT volumes and chest NCCT EAT volumes at different tube energies (120 KV vs. 100 KV) in Cohort 2. Bland-Altman plots (Left panel) and scatter plots (Right panel) of EAT volume between cardiac NCCT images acquired at 120 KV (referent) and chest NCCT-1.25 mm images, and between cardiac NCCT images acquired at 120 KV (referent) and chest NCCT-5 mm image acquired at 100 KV, respectively. Abbreviation: see Figure 1.

### 3.3 Impact of *Contrast enhancement on EAT from chest CT*

The effect of contrast agent on EAT volume quantification is summarized in Table 5 and Figure 5. Chest CECT-1.25mm scans produces EAT volumes that were consistently lower than those that were estimated in chest NCCT-1.25mm images (96.5±49.5 *vs*. 125.8±59.9 cm^3^, Δ%=-24.2±7.2%, *p* < 0.001). Performing the Bland-Altman analysis on chest CECT vs. chest NCCT EAT volume revealed a systemic underestimation of 29.4 (23.0, 35.7) cm^3^ if contrast enhancement was used. Similar results also appeared in the chest CECT-5mm EAT volume versus the chest NCCT-5mm EAT volume (91.6±48.8 *vs*. 125.8±59.9 cm^3^, Δ%=-25.0±7.7%, *p* < 0.001), and the Bland-Altman analysis showed a systematic underestimation of EAT volumes in contrast-enhanced scans by 27.4 (22.1, 32.7) cm^3^. Both chest CECT EAT volumes had strong correlations with corresponding chest NCCT EAT volumes, suggesting that inter-individual differences in EAT can be captured by imaging protocol, as long as acquisition protocol is consistent across all participants. Furthermore, the chest CECT EAT radiodensities were significantly higher (less negative) than the chest NCCT EAT radiodensities in both datasets [1.25 mm: (−72.5±7.0 HU for CECT *vs*. -73.5±7.4 HU for NCCT), *p* <0.05; 5 mm: (−69.6±6.6 HU for CECT *vs*. -70.8±6.5 HU for NCCT), *p* <0.05]. Additionally, the chest CECT-1.25 mm EAT volumes were higher than the chest CECT-5 mm EAT volumes (96.5±49.5 cm^3^ *vs*. 91.6±48.8cm^3^; Δ%=-6.5±6.7%, *p* < 0.001, see Table 5 & Figure 3C)

**Table 5.**
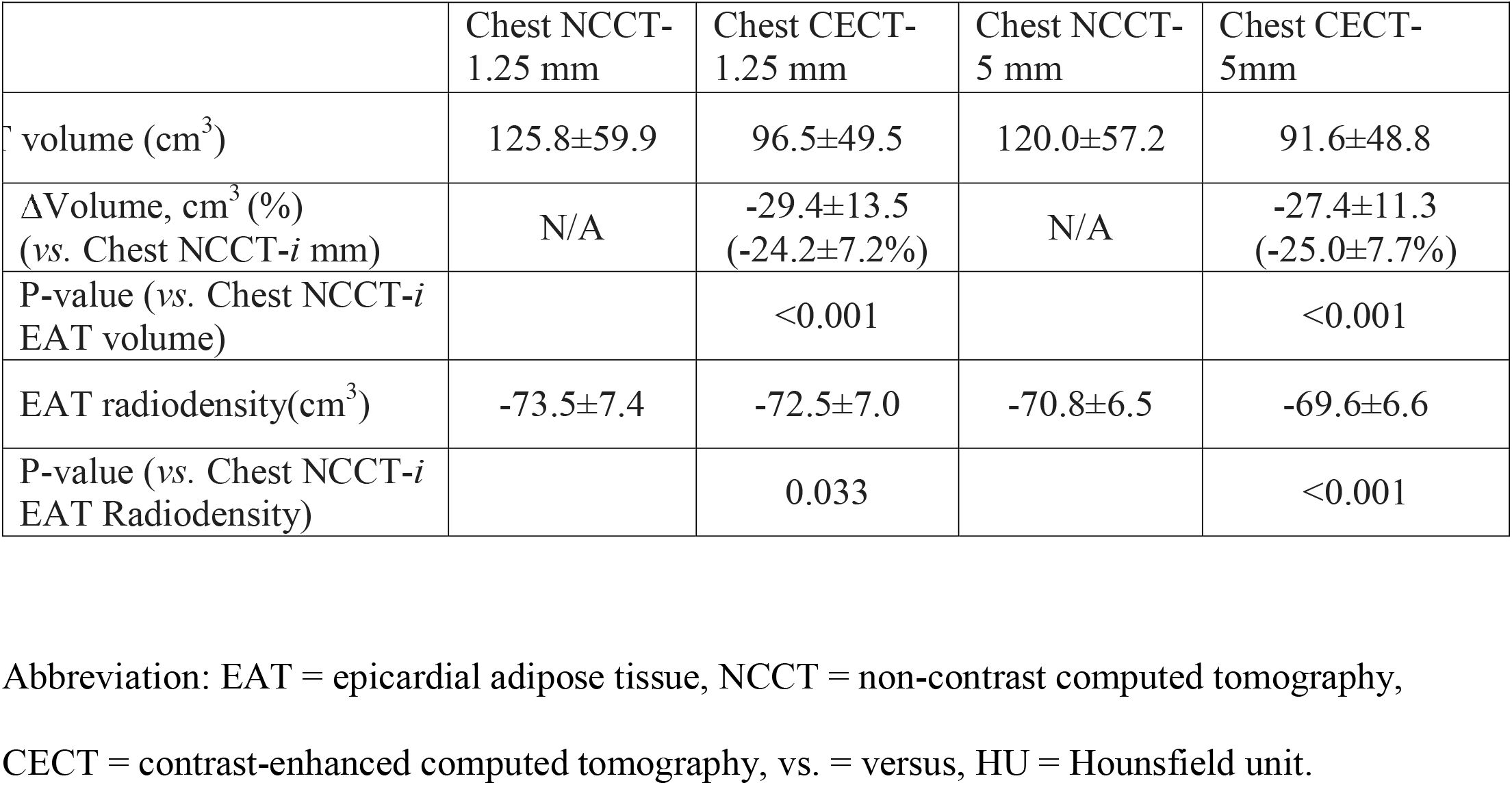
Impact of contrast enhancement on EAT volume in Cohort 3 (N=20)

**Figure 5.**
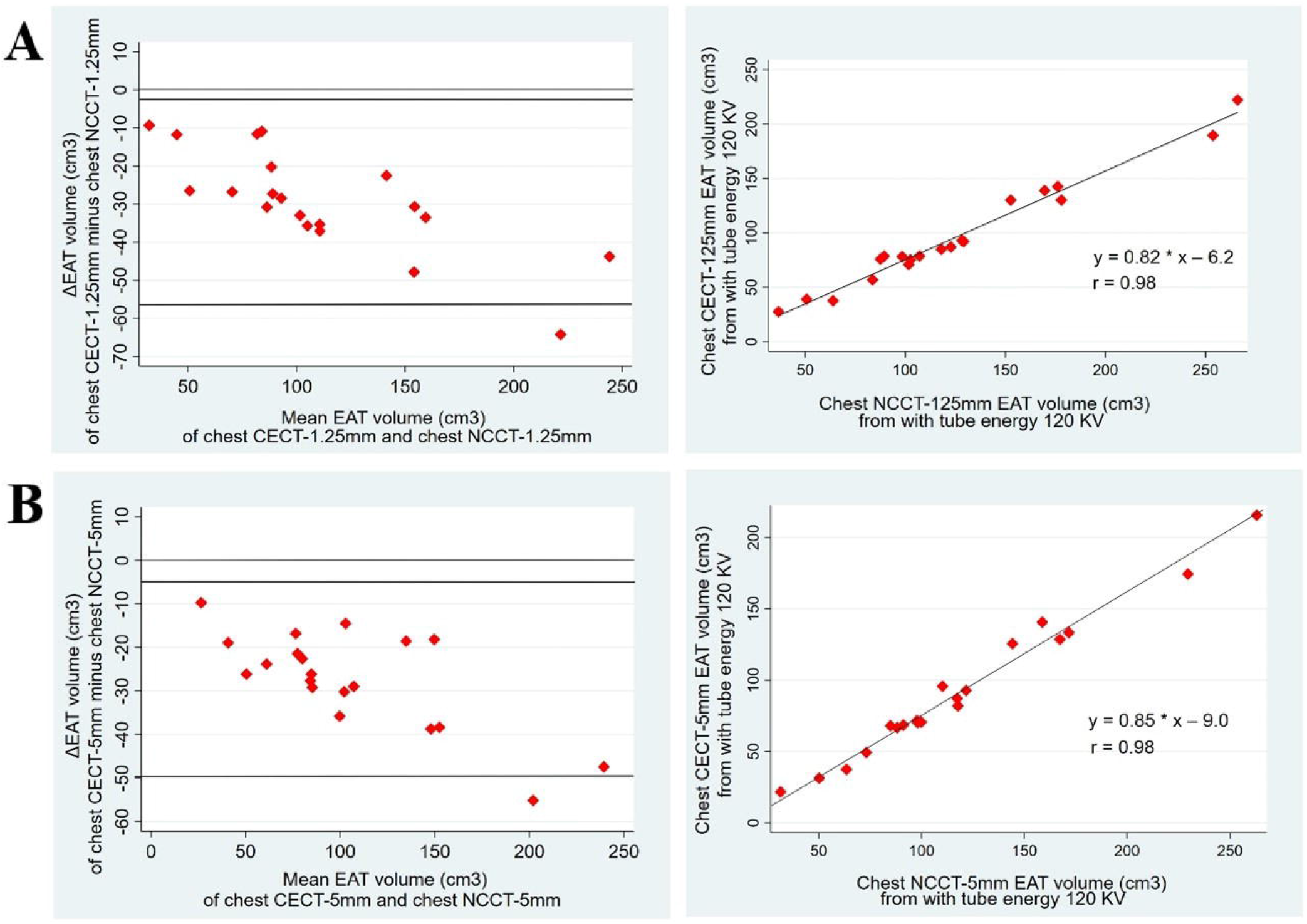
Effects of contrast agent on EAT volume quantification in paired chest NCCT and chest CECT images when other acquisition parameters remain constant. Bland-Altman plots (Left panel) and scatter plots (Right panel) of EAT volume between chest NCCT-1.25 mm images (referent) and chest CECT-1.25 mm images, and between chest NCCT-5 mm images (referent) and chest CECT-5 mm images, respectively. Abbreviation: see Figure 1.

## 4. Discussion

The main findings of the current study are: 1) the chest NCCT-1.25 mm image acquired with a tube energy 120 KV is an excellent alternative to the gold standard cardiac NCCT for producing accurate measurements of EAT volume; 2) similar to cardiac CT, contrast-enhanced chest CT scans underestimate EAT volumes; 3) thicker-sliced chest NCCT datasets (e.g., 5mm) underestimate EAT volumes, compared to thin-sliced (e.g., 1.25mm) for chest NCCT scans; 4) all chest CT derived EAT volumes are strongly correlated with the gold-standard EAT measurements.

To the best of our knowledge, this is the first study to systemically investigate the effects of acquisition and reconstruction parameters on EAT volume quantification in chest CT images, and to compare chest CT EAT volumes to those that were obtained using cardiac NCCT scans. Currently, cardiac CT scans are performed predominantly in patients with coronary artery disease.^21^ However, the clinical implications of EAT features should not only be evaluated in patients who undergo cardiac CT scan. Patients with chronic obstructive pulmonary disease, breast cancer, or lung cancer have a higher risk of coronary artery disease. ^8, 10^, ^22, 23 24, 25^ But these patients conventionally undergo chest, not cardiac, CT scans. Furthermore, chest CT scans have a much broader spectrum of clinical application, including disease of the lungs and the mediastinum, ect,^26^ with 11.6 million chest CT scans in 2006 in the United States.^9^ The fact that EAT volumes can be accurately estimated from chest CT scans will broaden the spectrum of EAT-associated research questions without additional radiation exposure and costly cardiac CT scanning.

### 4.1 EAT volume measured in cardiac NCCT vs chest NCCT with controlled tube energy

We found that the EAT volumes evaluated in chest NCCT-1.25 mm images acquired at a tube energy of 120 KV was almost identical to those derived from the gold standard cardiac NCCT acquired at 120 KV and with a 3-mm slice thickness. It is reasonable to assume this similarity is a consequence of additive ECG gating and slice thickness effects. As the 3-mm slice thickness images are not routinely reconstructed in chest CT scan, we were not able to directly compare the EAT volumes between the paired cardiac and chest CT dataset with the same slice thickness. We separately discussed the effect of ECG gating and slice thickness.

ECG gating technique has been widely used to mitigate motion artifacts in the assessment of coronary arteries, which move together with the left ventricle. Recently technical advances – e.g. increased speed of gantry rotation and pitch, application of dual-source multidetector CT scan – shortened imaging time and substantially improved temporal resolution of chest CT acquisition, further minimizing sensitivity to motion artifacts.^27^ The improved spatial resolution also enables more robust contouring of the pericardium in chest CT, and allows for segmentation of EAT. The ECG-gated CT images are typically acquired at the end diastole, while the non-ECG gated CT images can be attained continuously throughout the cardiac cycle. A recent CECT study that compared EAT volumes at systole and diastole concluded that the EAT volume is independent of the cardiac phase.^28^ Therefore, we believe that ECG gating has no significant impact on EAT volume quantification.

EAT volume is semi-automatically quantified based on the minimum unit of a voxel using the radiodensity threshold of -190, -30 HU. Although EAT predominantly contains fat, there are abundant small vessels, nerves, and immune cells^1^ which present much higher radiodensities +40 to +60 HU.^29^ Thicker slices, and thus larger voxel dimensions, increase the potential for partial volume errors, where a significant fraction of the voxel is occupied by material with different radiodensities, outside of the targeted range of -190, -30 HU. Consequently, the NCCT images with thicker slices, compared to the those with thinner slices, has a smaller percentage of voxels fall into the radiodensity range of -190, -30 HU. In the current study, voxel size in the chest NCCT-1.25 mm datasets [(0.60-0.65)* (0.60-0.65)*(0.7-1)] was close to that of the standard cardiac NCCT scans [(0.3-0.4)* (0.3-0.4)*1.5], but significantly smaller than the voxel size of the chest NCCT-5 mm images [(0.60-0.65)*(0.60-0.65)*5)]. This helps to explain that the chest NCCT-12.5 mm EAT volumes were similar to their cardiac counterparts but significantly larger than chest NCCT-5mm EAT volumes from the same participants. Furthermore, the EAT radiodensities of our chest NCCT-1.25mm scans were similar to those of cardiac NCCT but lower (i.e., more negative) than the EAT radiodensities derived from chest CT-5mm scans. This may have contributed to smaller EAT volumes in chest CT-5mm images without additional radiodensity threshold adjustments. Consistent with our findings, comparable coronary calcium scores have been reported in cardiac NCCT vs chest NCCT 1.25mm comparisons, while NCCT-5mm produced underestimates.^18^

In conclusion, we believe that ECG gating has no significant impact on EAT volume quantification, while the slice thickness has a significant contributing role for the similarity of EAT volume quantification between the chest NCCT-1.25 mm images and the standard cardiac NCCT image when other imaging parameters remain constant.

### 4.2 Tube energy and EAT volume

Compared to the standardized cardiac NCCT EAT volume with tube energy of 120 KV, which is similar to the chest NCCT-1.25 mm EAT volumes with tube energy of 120 KV, the chest NCCT-1.25 mm EAT volumes with tube energy of 100 KV were significantly overestimated. Consequently, a lower tube energy contributed to the overestimation of EAT volume. Additionally, we also observed that lowering the tube energy decreases (i.e., more negative) the EAT radiodensity. Tube energy has different impacts on tissues based on the tissue structure, and lower attenuation values (more negative) of pericardial fat at lower tube energy scan have been reported.^14, 30^ It is possible that lower EAT radiodensity values contributed to overestimation of EAT volumes, and different radiodensity thresholds are optimal for EAT volume quantification at different tube energies. Furthermore, lower tube energy reduces image signal to noise ratio, which may also impact the accuracy of EAT estimation.^31^ Similar to our current results, Marwan et al found a significant overestimation of the cardiac NCCT EAT volume using lower tube energy.^31^ The slightly lower overestimation of EAT volume in chest NCCT-5 mm acquisitions at 100 KV are most likely the result of EAT underestimation due to thicker slices.

### 4.3 Contrast agent vs. EAT volume

The current study is the first to confirm that the chest CECT EAT volumes were significantly smaller than paired chest NCCT EAT volumes, regardless of slice thicknesses (i.e., 5 mm and 1.25 mm). In our previous study, we have shown that EAT volumes derived from cardiac CECT datasets were significantly smaller than those derived from the cardiac NCCT datasets when using the standard radiodensity threshold (−190, -30) HU.^17^ Taken together, our results indicate that adding a contrast agent increases the EAT radiodensity (less negative) in both cardiac and chest CT scans, and threshold adjustment might be necessary to correct contrast-induced biases.^17^

### 4.4 Correlations of EAT volume between chest CT images and cardiac NCCT image

We observed strong correlations between chest NCCT EAT volumes and those from the standard cardiac NCCT images, as well as between chest CT-5mm EAT volumes and those from chest CT-1.25 mm images, and between chest NCCT EAT volumes and those from chest CECT images. Consequently, even though only the chest NCCT-1.25 mm images produced similar absolute EAT volumes to standard cardiac NCCT scans, it is likely that EAT volumes from chest CT scans, regardless of acquisition parameters, carry similar prognostic values as those of conventional cardiac CT measures, as long as the same imaging acquisition protocol is applied to all subjects in a study. On the other hand, a mixed use of chest CT dataset, with inconsistent tube energies, slice thicknesses, and use of contrast agent, is not recommended.

### 4.5 Limitations

First, the investigation of the ECG gate should be ideally between cardiac NCCT and chest NCCT with the same slice thickness (i.e., 3 mm). However, the chest NCCT-3mm dataset was not available, because chest CT images are not conventionally reconstructed with 3-mm slice thickness. Second, the investigation of the tube energy would ideally be explored in paired chest CT datasets with different tube energies, which were not available in this retrospective study. However, in Cohort 2, cardiac NCCT EAT volume acquired at 120 KV was an excellent alternative of the chest NCCT 1.25 mm EAT volume acquired at 120 KV. Third, more various slice thicknesses should be explored. However, the slice thicknesses 1.25 mm and 5 mm are the most commonly used in clinical practice, and we believe they are sufficiently representable. And we believe that, as long as other chest CT images with different slice thicknesses have the voxel size substantially close to that of the cardiac NCCT image, they might generate a similar EAT volumes, if other imaging acquisitions and reconstruction parameters are held constant. Finally, the impact of interslice gap on EAT volume quantification should be further investigated in future studies.

## 5. Conclusion

The chest non-contrast CT image with a slice thickness of 1.25 mm at the tube energy of 120 KV is an excellent alternative of the standard cardiac non-contrast CT image to measure EAT volume. All chest CT EAT volumes were strongly corelated with cardiac counterparts.

## Data Availability

All data are available based on the reasonable requests.

## Acknowledgments

None

